# A 6-mRNA host response whole-blood classifier trained on pre-pandemic data accurately predicts severity in COVID-19 and other acute viral infections

**DOI:** 10.1101/2020.12.07.20230235

**Authors:** Ljubomir Buturovic, Hong Zheng, Benjamin Tang, Kevin Lai, Win Sen Kuan, Mark Gillett, Rahul Santram, Maryam Shojaei, Raquel Almansa, Jose Ángel Nieto, Sonsoles Muñoz, Carmen Herrero, Nikolaos Antonakos, Panayiotis Koufargyris, Marina Kontogiorgi, Georgia Damoraki, Oliver Liesenfeld, James Wacker, Uros Midic, Roland Luethy, David Rawling, Melissa Remmel, Sabrina Coyle, Yiran E. Liu, Aditya M Rao, Denis Dermadi, Jiaying Toh, Lara Murphy Jones, Michele Donato, Purvesh Khatri, Evangelos J. Giamarellos-Bourboulis, Timothy E Sweeney

## Abstract

**Background:** Determining the severity of COVID-19 remains an unmet medical need. Our objective was to develop a blood-based host-gene-expression classifier for the severity of viral infections and validate it in independent data, including COVID-19.

**Methods:** We developed the classifier for the severity of viral infections and validated it in multiple viral infection settings including COVID-19. We used training data (N=705) from 21 retrospective transcriptomic clinical studies of influenza and other viral illnesses looking at a preselected panel of host immune response messenger RNAs.

**Results:** We selected 6 host RNAs and trained logistic regression classifier with a cross-validation area under curve of 0.90 for predicting 30-day mortality in viral illnesses. Next, in 1,417 samples across 21 independent retrospective cohorts the locked 6-RNA classifier had an area under curve of 0.91 for discriminating patients with severe vs. non-severe infection. Next, in independent cohorts of prospectively (N=97) and retrospectively (N=100) enrolled patients with confirmed COVID-19, the classifier had an area under curve of 0.89 and 0.87, respectively, for identifying patients with severe respiratory failure or 30-day mortality. Finally, we developed a loop-mediated isothermal gene expression assay for the 6-messenger-RNA panel to facilitate implementation as a rapid assay.

**Conclusions:** With further study, the classifier could assist in the risk assessment of COVID-19 and other acute viral infections patients to determine severity and level of care, thereby improving patient management and reducing healthcare burden.

## Background

The emergence of the SARS-coronavirus 2 (SARS-CoV-2), causative agent of COVID-19, and its rapid pandemic spread has led to a global health crisis with more than 54 million cases and more than 1 million deaths to date (*1*). COVID-19 presents with a spectrum of clinical phenotypes, with most patients exhibiting mild-to-moderate symptoms, and 20% progressing to severe or critical disease, typically within a week (*2-6*). Severe cases are often characterized by acute respiratory failure requiring mechanical ventilation and sometimes progressing to ARDS and death (*7*). Illness severity and development of ARDS are associated with older age and underlying medical conditions (*3*).

Yet, despite the rapid progress in developing diagnostics for SARS-CoV-2 infection, existing prognostic markers ranging from clinical data to biomarkers and immunopathological findings have proven unable to identify which patients are likely to progress to severe disease (*8*). Poor risk stratification means that front-line providers may be unable to determine which patients might be safe to quarantine and convalesce at home, and which need close monitoring. Early identification of severity along with monitoring of immune status may also prove important for selection of treatments such as corticosteroids, intravenous immunoglobulin, or selective cytokine blockade (*9-11*).

A host of lab values, including neutrophilia, lymphocyte counts, CD3 and CD4 T-cell counts, interleukin-6 and -8, lactate dehydrogenase, D-dimer, AST, prealbumin, creatinine, glucose, low-density lipoprotein, serum ferritin, and prothrombin time rather than viral factors have been associated with higher risk of severe disease and ARDS (*3, 12, 13*). While combining multiple weak markers through machine learning (ML) has a potential to increase test discrimination and clinical utility, applications of ML to date have led to serious overfitting and lack of clinical adoption (*14*). The failure of such models arises both from a lack of clinical heterogeneity in training, and from the pragmatic nature of the variable selection, which uses existing lab tests which may not be ideal for the task. Furthermore, a number of the lab markers are late indicators of severity since by the time they become abnormal, patient is already very sick.

The host immune response represented in the whole blood transcriptome has been repeatedly shown to diagnose presence, type, and severity of infections (*15-19*). By leveraging clinical, biological, and technical heterogeneity across multiple independent datasets, we have previously identified a conserved host response to respiratory viral infections (*16*) that is distinct from bacterial infections (*15-17*) and can identify asymptomatic infection. Recently, we have demonstrated that this conserved host response to viral infections is strongly associated with severity of outcome, including in patients infected with SARS-CoV-2, chikungunya, and Ebola (20). We have also demonstrated that conserved host immune response to infection can be an accurate prognostic marker of risk of 30-day mortality in patients with infectious diseases (*18*). Most importantly, we have demonstrated that accounting for biological, clinical, and technical heterogeneity identifies more generalizable robust host response-based signatures that can be rapidly translated on a targeted platform (*19*).

Based on these previous results that there is a shared blood host-immune response-based mRNA prognostic signature among patients with acute viral infections, we hypothesized that a parsimonious, clinically translatable gene signature for predicting outcome in patients with viral infection can be identified. We tested this hypothesis by integrating 21 independent data sets with 705 peripheral blood transcriptome profiles from patients with acute viral infections and identified a 6-mRNA host-response-based signature for mortality prediction across these multiple viral datasets. Next, we validated the locked model in another 21 independent retrospective cohorts of 1,417 blood transcriptome profiles of patients with a variety of viral infections (non-COVID). Finally, we validated our 6-mRNA model in independent prospectively and retrospectively collected cohorts of patients with COVID-19, demonstrating its ability to predict outcomes despite having been entirely trained using non-COVID data. Our results suggest the conserved host response to acute viral infection can be used to predict its outcome. Finally, we showed validity of a rapid isothermal version of the 6-mRNA host-response-signature which is being further developed into a rapid molecular test (CoVerity™) to assist in improving management of patients with COVID-19 and other acute viral infections.

## Materials and Methods

### Data collection, curation, and sample labeling

We searched public repositories (National Center for Biotechnology Information Gene Expression Omnibus and European Bioinformatics Institute ArrayExpress) for studies of typical acute infection with mortality data present. After removal of pediatric and entirely non-viral datasets, we identified 17 microarray or RNAseq peripheral blood acute infection studies composed of samples from 1,861 adult patients with either 28-day or 30-day mortality information (**Figure 1** and **Table 1**). We processed and co-normalized these datasets as described previously (*19*).

**Table 1.** Characteristics of viral infection studies used for training. *COPD, chronic pulmonary obstruction disorder; ** ICU, intensive care unit; ***TB, tuberculosis; ****CAP, community-acquired penumonia

| Study identifier | First author or PI | Study description | Timing of sample collection | N (survivors/non-survivors) | Age (Median, IQR) | Male (n (%)) | Country | Platform |
| --- | --- | --- | --- | --- | --- | --- | --- | --- |
| E-MEXP-3589 | Almansa | Patients hospitalized with COPD* exacerbation | Hospital/ICU admission | 5 (5/0) | Unk. | 5(100) | Spain | Agilent |
| E-MTAB-1548 | Almansa | Surgical patients with sepsis (EXPRESS) | Average post-operation day 4 | 3 (3/0) | 78.0 (71.5-79.5) | 3(100) | Spain | Agilent |
| E-MEXP-3162 | Van de Weg | Uncomplicated dengue | Within 48h of onset | 21 (21/0) | Unk. | Unk. | Indonesia | Affymetrix |
| GSE13015 (GPL6102) | Pankla | Sepsis, many cases from burkholderia | Within 48 h of diagnosis | 3 (2/1) | 54.0 (46.0-55.5) | 1(33) | Thailand | Illumina |
| GSE13015 (GPL6947) |  |  |  | 2 (2/0) | 64.5 (56.2-72.8) | 1(50) |  | Illumina |
| GSE21802 | Bermejo-Martin | Pandemic H1N1 in ICU** | Within 48 h of ICU admission | 6 (5/1) | Unk. | Unk. | Canada | Illumina |
| GSE22098 | Berry | Patients with active TB*** and other inflammatory and infectious diseases | At admission | 39 (39/0) | 31.0 (19.0-47.0) | 6(15) | UK, South Africa | Illumina |
| GSE27131 | Berdal | Severe H1N1 influenza | Admission to ICU | 3 (2/1) | 38.0 (31.5-46.0) | 3(100) | Norway | Affymetrix |
| GSE28991 | Naim | Acute dengue fever | Within 72h of onset | 11 (11/0) | Unk. | Unk. | Singapore | Illumina |
| GSE32707 | Dolinay | Critically ill patients in ICU (Sepsis, SIRS and/or ARDS) | Admission to ICU | 7 (5/2) | 45.0 (39.0-50.5) | 4(57) | USA | Illumina |
| GSE40012 | Parnell | Bacterial or influenza A pneumonia or SIRS | Admission to ICU | 11 (11/0) | Unk. | 4(36) | Australia | Illumina |
| GSE54514 | Parnell | Sepsis patients in ICU | Admission to ICU | 2 (2/0) | 62.5 (60.2-64.8) | 1(50) | Australia | Illumina |
| GSE51808 | Kwissa | Acute dengue fever | 1-8 days after onset | 28 (28/0) |  | Unk. | Thailand | Affymetrix |
| GSE60244 | Suarez | Lower respiratory tract infections | Within 24 h of admission | 62 (62/0) | 59.0 (50.0-74.5) | 24(39) | USA | Illumina |
| GSE65682 | Scicluna | Suspected but negative for CAP**** | Within 24 h of ICU admission | 9 (7/2) | 67.0 (63.0-73.0) | 7(78) | Netherlands | Affymetrix |
| GSE68310 | Zhai | Outpatients with acute respiratory viral infections | Within 48h of onset | 75 (75/0) | 21.0 (20.4-22.3) | 34(45) | USA | Illumina |
| GSE82050 | Tang | Moderate and severe influenza infection | Within 24 h of admission | 17 (17/0) | 55.0 (45.0-72.0) | Unk. | Germany | Agilent |
| GSE95233 | Venet | Septic shock patients in ICU | Admission to ICU | 7 (5/2) | 47.0 (42.0-65.0) | 5(71) | France | Affymetrix |
| Australia / WIMR | Tang | Community or hospital clinics with influenza-like illness | At presentation | 332 (321/11) | 48.0 (32.0-63.5) | 129(39) | Australia | Nanostring |
| Stanford ICU databank | Rogers | Suspected sepsis with ARDS risk factors | Admission to ICU | 8 (6/2) | 62.0 (55.5-67.2) | 4(50) | USA | Nanostring |
| PROMPT | Giamarellos-Bourboulis | Suspected infection with 2+ SIRS | Admission to ED | 1 (1/0) | 78.0 | 0(0) | Greece | Nanostring |
| PREVISE | Herrero | Outpatient urgent care with suspected CAP | At presentation | 53 (52/1) | 78.0 (66.0-87.0) | 33(62) | Spain | Nanostring |

**Figure 1.**
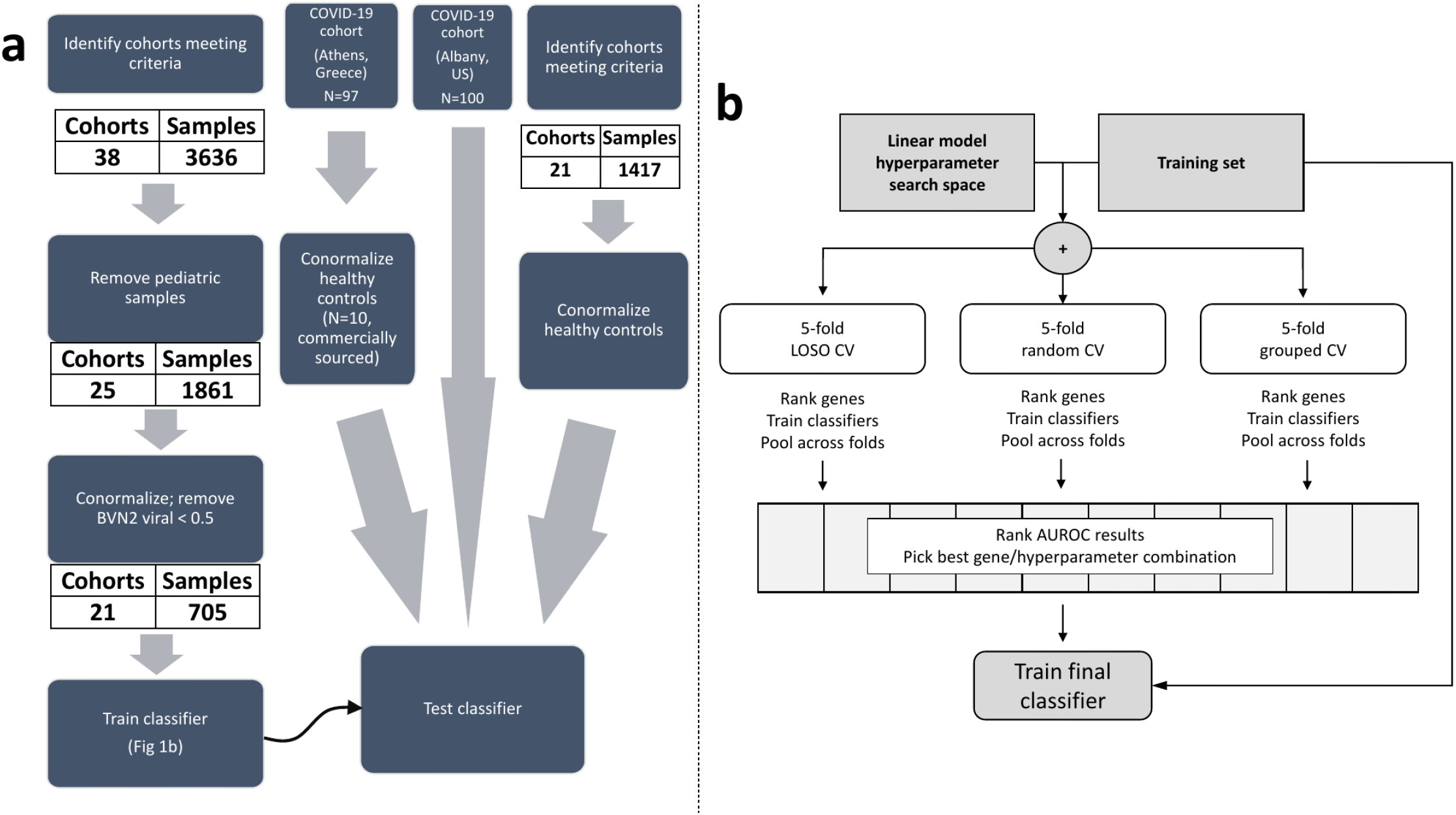
Study flow. (a) Clinical data flows for training and testing. (b) Machine learning worfklow used to develop and validate the 6-mRNA viral severity classifier. LOSO = Leave-One-Study-Out. CV = cross-validation. AUROC = Area Under ROC curve.

The number of cases with clinically adjudicated viral infection and known mortality outcome among the public samples was too low for robust modeling. Thus, to increase the number of training samples, we assigned viral infection status using a previously developed gene-expression-based bacterial/viral classifier, whose accuracy approaches that of clinical adjudication. Specifically, we utilized an updated version of our previously described neural network-based classifier for diagnosis of bacterial vs. viral infections called ‘Inflammatix Bacterial-Viral Noninfected version 2’ (IMX-BVN-2), (*18*). The rationale is that this method would increase the number of mortality samples with viral infection, without introducing many false positives. For all samples, we applied IMX-BVN-2 to assign a probability of bacterial or viral infection and retained samples for which viral probability according to IMX-BVN-2 was ≥0.5. We refer to this assessment of viral infection as computer-aided adjudication. Out of 1,861 samples, we found 311 samples which had IMX-BVN-2 probability of viral infection ≥0.5, of which 9 patients died within 30-day period.

In addition to this public microarray/RNAseq data, we included 394 samples across 4 independent cohorts (*19*) that were profiled using NanoString nCounter, of which 14 patients died (**Table 1**). Thus, overall we included 705 blood samples across 21 independent studies from patients with computer aided-adjudication of viral infection and short-term mortality outcome. Importantly, none of these patients had SARS-CoV-2 infection as they were all enrolled prior to November 2019.

### Selection of variables for classifier development

We preselected 29 mRNAs from which to develop the classifier for several biological and practical reasons. Biologically, the 29 mRNAs are composed of an 11-gene set for predicting 30-day mortality in critically ill patients and a repeatedly validated 18-gene set that can identify viral vs bacterial or noninfectious inflammation (*17-19*). Thus, we hypothesized that if a generalizable viral severity signature were possible, we likely had appropriate (and pre-vetted) variables here. By limiting our input variables, we also lowered our risk of overfitting to the training data. From a practical perspective, first, we are developing a point-of-care diagnostic platform for measuring these 29 genes in less than 30 minutes. Hence, a classifier developed using a subset of these 29 genes would allow us to develop a rapid point-of-care test on our existing platform. Second, 4 of the 21 cohorts included in the training were Inflammatix studies that profiled these 29 genes using NanoString nCounter and therefore for those studies this was the only mRNA expression data available.

### Development of a classifier using machine learning

We analyzed the 705 viral samples using cross-validation (CV) for ranking and selecting machine learning classifiers. We explored three variants of cross-validation: (1) 5-fold random CV, (2) 5-fold grouped CV, where each fold comprises multiple studies, and each study is assigned to exactly one CV fold, and (3) leave-one-study-out (LOSO), where each study forms a CV fold. We included non-random CV variants because we recently demonstrated that the leave-one-study-out cross-validation may reduce overfitting during training and produce more robust classifiers, for certain datasets (*19*). The hyperparameter search space was based on machine learning best practices and our previous results in model optimization in infectious disease diagnostics (*21*). For rapid turnaround and to reduce overfitting, we only investigated linear classifiers (support vector machine with linear kernel, logistic regression, and multi-layer perceptron with linear activation function) and limited the number of hyperparameter configurations we searched to 1000 per classifier. Finally, to ensure a parsimonious signature for translation to a rapid molecular assay, we limited the number of genes in the final model to six. To select the six genes, we applied forward selection and univariate feature ranking. We followed best practices to avoid overfitting in the gene selection process (*22, 23*).

We performed cross-validations for each of the hyperparameter configurations. Within each fold, we sorted the absolute value of the genes’ Pearson correlation with class label (survived/died). We then trained a classifier using the six top-ranked genes and applied it to the left-out fold. The predicted probabilities from the folds were pooled, and the Area Under a Receiver Operating Characteristic (AUROC) curve over the pooled cross-validation probabilities was used as a metric to rank classification models. The final ranking of genes was determined using average ranking across the CV folds. Once the best-ranking model hyperparameters were selected and the final list of six genes was established, the final model was trained using the entire training set and the ‘locked’ hyperparameters. The corresponding model weights were locked and the final classifier was then tested in an independent prospective cohort of patients with COVID-19, and in independent retrospective cohort of patients with viral infections without COVID-19.

### Retrospective non-COVID-19 patient cohort

We selected a subset of samples from our previously described database of 34 independent cohorts derived from whole blood or peripheral blood mononuclear cells (PBMCs) (*20*). From this database we removed all samples that were used in our analysis for identifying the 6-gene signature, leaving 1,417 samples across 21 independent cohorts (**Supplementary Table 1**). The samples in these datasets represented the biological and clinical heterogeneity observed in the real-world patient population, including healthy controls and patients infected with 16 different viruses with severity ranging from asymptomatic to fatal viral infection over a broad age range (<12 months to 73 years) (**Figure 1A** and **Supplementary Table 1**). Notably, the samples were from patients enrolled across 10 different countries representing diverse genetic backgrounds of patients and viruses. Finally, we included technical heterogeneity in our analysis as these datasets were profiled using microarray from different manufacturers.

We renormalized all microarray datasets using standard methods when raw data were available from the Gene Expression Omnibus database. We applied Guanine Cytosine Robust Multiarray Average to arrays with mismatch probes for Affymetrix arrays. We used normal-exponential background correction followed by quantile normalization for Illumina, Agilent, GE, and other commercial arrays. We did not renormalize custom arrays and used preprocessed data as made publicly available by the study authors. We mapped microarray probes in each dataset to Entrez Gene identifiers (IDs) to facilitate integrated analysis. If a probe matched more than one gene, we expanded the expression data for that probe to add one record for each gene. When multiple probes mapped to the same gene within a dataset, we applied a fixed-effect model. Within a dataset, cohorts assayed with different microarray types were treated as independent.

### Standardized severity assignment for retrospective non-COVID-19 patient samples

We used standardized severity for each of the 1,417 samples as described before (*20*). Briefly, for each dataset, we used the sample phenotypes as defined in the original publication. We manually assigned a severity category to each sample based on the cohort description for each dataset in the original publication as follows: (1) healthy controls – asymptomatic, uninfected healthy individuals, (2) asymptomatic or convalescents – afebrile asymptomatic individuals who tested positive for a virus or those fully recovered from a viral infection with completely resolved symptoms, (3) mild – symptomatic individuals with viral infection that were either managed as outpatient or discharged from the emergency department (ED), (4) moderate – symptomatic individuals with viral infection who were admitted to the general wards and did not require supplemental oxygen, (5) serious - symptomatic individuals with viral infection who were described as ‘severe’ by original authors, admitted to general wards with supplemental oxygen, or admitted to the intensive care unit (ICU) without requiring mechanical ventilation or inotropic support, (6) critical - symptomatic individuals with viral infection who were on mechanical ventilation in the ICU or were diagnosed with acute respiratory distress syndrome (ARDS), septic shock, or multiorgan dysfunction syndrome, and (7) fatal – patients with viral infection who died in the ICU.

For datasets that did not provide sample-level severity data (GSE101702, GSE38900, GSE103842, GSE66099, GSE77087), we assigned severity categories as follows. We categorized all samples in a dataset as “moderate” when either (1) >70% of patients were admitted to the general wards as opposed to discharged from the ED, (2) <20% of patients admitted to the general wards required supplemental oxygen, or (3) patients were admitted to the general wards and categorized as ‘mild’ or ‘moderate’ by the original authors. We categorized all samples in a dataset as “severe” when >20% of patients had either (1) been admitted to the general wards and categorized as ‘severe’ by original authors, (2) required supplemental oxygen, or (3) required ICU admission without mechanical ventilation.

### Retrospective COVID-19 patient cohort

We used COVID-19 samples (N=100) from GSE 157103 (*24*). Briefly, the dataset includes RNA-Seq expression data for adult patients hospitalized for suspected COVID-19 in April 2020, in Albany Medical Center (Albany, NY, United States). We applied the 6-mRNA classifier to the gene expression data for participants who tested positive for COVID-19. The expression values were estimated by “RNA-Seq by Expectation Minimization” algorithm by the study authors. We used the binary “mechanical-ventilation” status (yes/no) provided by authors as indicator of the disease severity.

### Prospective COVID-19 patient cohort

Blood samples were collected between March and April 2020 from three study sites participating in the Hellenic Sepsis Study Group (www.sepsis.gr). The studies were conducted following approvals for the collection of biomaterial for transcriptomic analysis for patients with lower respiratory tract infections provided by the Ethics Committees of the participating hospitals. Participants were adults with written informed consent provided by themselves or by first-degree relatives in the case of patients unable to consent, with molecular detection of SARS-CoV-2 in respiratory secretions and radiological evidence of lower respiratory tract involvement. PAXgene® Blood RNA tubes were drawn within the first 24 hours from admission along with other standard laboratory parameters. Data collection included demographic information, clinical scores (SOFA, APACHE II), laboratory results, length of stay and clinical outcomes. Patients were followed up daily for 30 days; severe disease was defined as respiratory failure (PaO2/FiO2 ratio less than 150 requiring mechanical ventilation) or death. PAXgene Blood RNA samples were shipped to Inflammatix, where RNA was extracted and processed using NanoString nCounter®, as previously described (*19*). The 6-mRNA scores were calculated after locking the classifier weights.

### Healthy controls

We acquired five whole blood samples from healthy controls through a commercial vendor (BioIVT). The individuals were non-febrile and verbally screened to confirm no signs or symptoms of infection were present within 3 days prior to sample collection. They were also verbally screened to confirm that they were not currently undergoing antibiotic treatment and had not taken antibiotics within 3 days prior to sample collection. Further, all samples were shown to be negative for HIV, West Nile, Hepatitis B, and Hepatitis C by molecular or antibody-based testing. Samples were collected in PAXgene Blood RNA tubes and treated per the manufacturer’s protocol. Samples were stored and transported at -80C.

### Rapid isothermal assay

Our goal was to create a rapid assay, and isothermal reactions run much faster than traditional qPCR. Thus, loop-mediated isothermal gene expression (LAMP) assays were designed to span exon junctions, and at least three core (FIP/BIP/F3/B3) solutions meeting these design criteria were identified for each marker and evaluated for successful amplification of cDNA and exclusion of gDNA. Where available, loop primers (LF/LB) were subsequently identified for best core solutions to generate a complete primer set. Solutions were down-selected based on efficient amplification of cDNA and RNA, selectivity against gDNA, and the presence of single, homogenous melt peaks. The final primer sets are attached as **Supplementary Table 2**.

We designed an analytical validation panel of 61 blood samples from patients in multiple infection classes, including healthy, bacterial or viral. A subset of samples from patients with bacterial or viral infection came from patients with an infection that had progressed to sepsis. Whole blood samples were collected in PAXgene Blood RNA stabilization vacutainers, which preserve the integrity of the host mRNA expression profile at the time of draw. Total RNA was extracted from a 1.5 mL aliquot of each stabilized blood sample using a modified version of the Agencourt RNAdvance Blood kit and protocol. RNA was heat treated at 55°C for 5 min then snap-cooled prior to quantitation. Total RNA material was distributed evenly across LAMP reactions measuring the five markers in triplicate. LAMP assays were carried out using a modified version of the protocol recommended by Optigene Ltd, and performed on a QuantStudio 6 Real-Time PCR System.

### Statistical Analyses

Analyses were performed in R version 3 and Python version 3.6. The area under the receiver operating characteristic curve (AUROC) was chosen as the primary metric for model evaluation since it provides a general measure of diagnostic test quality without depending on prevalence or having to choose a specific cutoff point.

All validation dataset analyses use the locked 6-mRNA logistic regression output, i.e. predicted probabilities. AUROCs for additional markers (**Table 3**) are calculated from the available data for each marker. For the logistic regression model that includes the 6-mRNA predicted probabilities along with other markers as predictor variables, conditional multiple imputation was used for values to ensure model convergence. Since AUROC may fail to detect poor calibration on validation data (since subject rankings may still hold), we also demonstrated that a cutoff chosen from training data maintains good sensitivity and specificity in validation data even before recalibration. Due to the relatively small sample size, we made inter-group comparisons without assumptions of normality where possible (Kruskal-Wallis rank sum or Mann-Whitney U test). Medians and interquartile ranges are given for continuous variables.

**Table 2.** Demographics, severity scores, and severity markers for the prospective COVID-19 cohort, overall and split by mortality. P-values correspond to Mann-Whitney tests for difference of means and chi-square tests for difference of proportions between the survival and mortality groups. Unless indicated otherwise, numbers shown are median [IQR].

| Variable | Overall | Death | Survival | P value |
| --- | --- | --- | --- | --- |
| N | 97 | 16 | 81 |  |
| Age years | 62 [52, 72.25] | 68.50 [62.75, 84.25] | 60.00 [50.75, 70.25] | 0.003 |
| Gender = Male (%) | 68 (70.1) | 12 ( 75.0) | 56 (69.1) | 0.865 |
| White blood cells /mm3 | 6770 [5145, 10227.50] | 8540.00 [5542.50, 12510.00] | 6480.00 [5145.00, 9622.50] | 0.275 |
| Neutrophils (%) | 78.10 [68.35, 86.60] | 88.95 [86.40, 93.03] | 77.09 [65.22, 83.75] | <0.001 |
| Lymphocytes (%) | 12.70 [7.20, 21.15] | 6.70 [3.65, 9.65] | 14.03 [9.00, 22.42] | <0.001 |
| Platelets /mm3 | 215000 [172900, 266000] | 249050 [180750, 298000] | 214000 [172600, 260800] | 0.176 |
| D-dimer ng/ml | 977.90 [476.25, 2560.00] | 4480.00 [2440.00, 13161.50] | 850.00 [437.50, 1947.50] | <0.001 |
| CRP mg/l | 107.00 [31.60, 222.50] | 224.75 [142.89, 260.75] | 79.10 [28.80, 202.00] | 0.002 |
| SOFA score | 3.00 [1.00, 6.00] | 5.50 [4.00, 6.25] | 2 [1, 6] | 0.006 |
| APACHE II | 7.00 [5.00, 11.00] | 11.00 [8.00, 13.50] | 7 [4, 9] | 0.001 |
| Length of hospital stay | 13.00 [11.00, 20.00] | 13 [8.75, 17.25] | 13 [11, 20] | 0.410 |
| Severe respiratory failure (%) | 50 (51.5) | 16 (100.0) | 34 (42.0) | <0.001 |

**Table 3.**
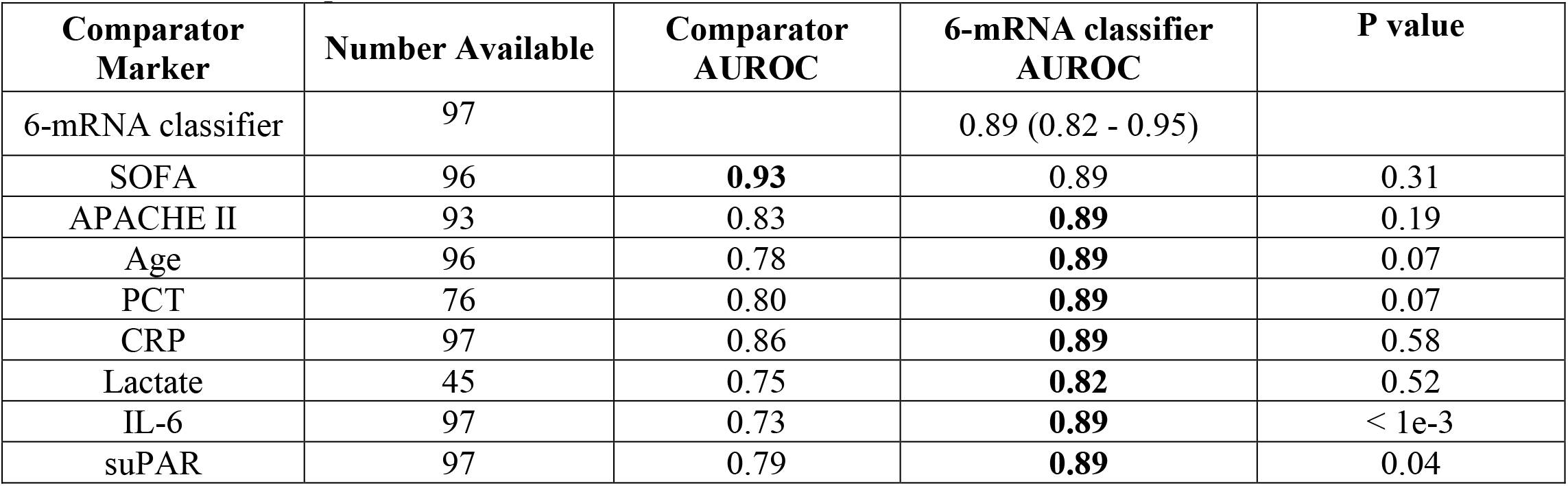
Prognostic power of the 6-mRNA signature classifier and comparator scores and markers in the independent prospective COVID-19 cohort. Shown are AUROCs for non-missing data, plus 95% CI. The final column is a ‘fair’ assessment of the 6-mRNA signature classifier, i.e. the performance on the subset of patients that was available to the comparator. **Table 3a.** Prognostic power for predicting severe respiratory failure. Bold font indicates predictor with higher AUROC, which in nearly all cases is the 6-mRNA classifier. The P value column corresponds to DeLong test for difference between paired ROC curves.

**Table 3b.** Prognostic power for predicting mortality. Bold font indicates predictor with the higher AUROC. The P value column corresponds to DeLong test for difference between paired ROC curves.

| Comparator Marker | Number Available | Comparator AUROC | 6-mRNA classifier AUROC | P value |
| --- | --- | --- | --- | --- |
| 6-mRNA classifier | 97 |  | 0.78 (0.64 - 0.92) |  |
| SOFA | 96 | 0.72 | **0.78** | 0.43 |
| APACHE II | 93 | 0.76 | **0.77** | 0.84 |
| Age | 96 | 0.74 | **0.78** | 0.68 |
| PCT | 76 | 0.73 | **0.77** | 0.63 |
| CRP | 97 | 0.74 | **0.78** | 0.62 |
| Lactate | 45 | 0.78 | **0.80** | 0.84 |
| IL-6 | 97 | 0.57 | **0.78** | 0.003 |
| suPAR | 97 | 0.74 | **0.78** | 0.71 |

## Results

We first identified 21 studies (*25-40*) with 705 patients with viral infections (no patient with SARS-CoV-2) based on computer-aided adjudication and available outcomes data (see **Methods**; **Figure 1** and **Table 1**). These studies included a broad spectrum of clinical, biological, and technical heterogeneity as they profiled blood samples from viral infections from 14 countries using mRNA profiling platforms from four manufacturers (Affymetrix, Agilent, Illumina, Nanostring). Within each dataset, the number of patients who died were very low (two or less for all but one study), meaning traditional approaches for biomarker discovery that rely on a single cohort with sufficient sample size would not have been effective. However, there were sufficient cases (23 deaths within 30 days of sample collection) across these 705 patients. Sample size analysis using pmsampsize package (41) suggested minimal sample size of 450 patients with 18 cases, confirming the adequacy of the pooled dataset. Our previously described approaches for integrating independent datasets and leveraging heterogeneity allowed us to learn across the whole pooled dataset (*19, 42, 43*). Visualization of the 705 conormalized samples using all genes present across the studies using t-stochastic neighbor embedding (t-SNE), showed that there was no clear separation between the samples from patients who died and those who survived (**Figure 2a**).

**Figure 2.**
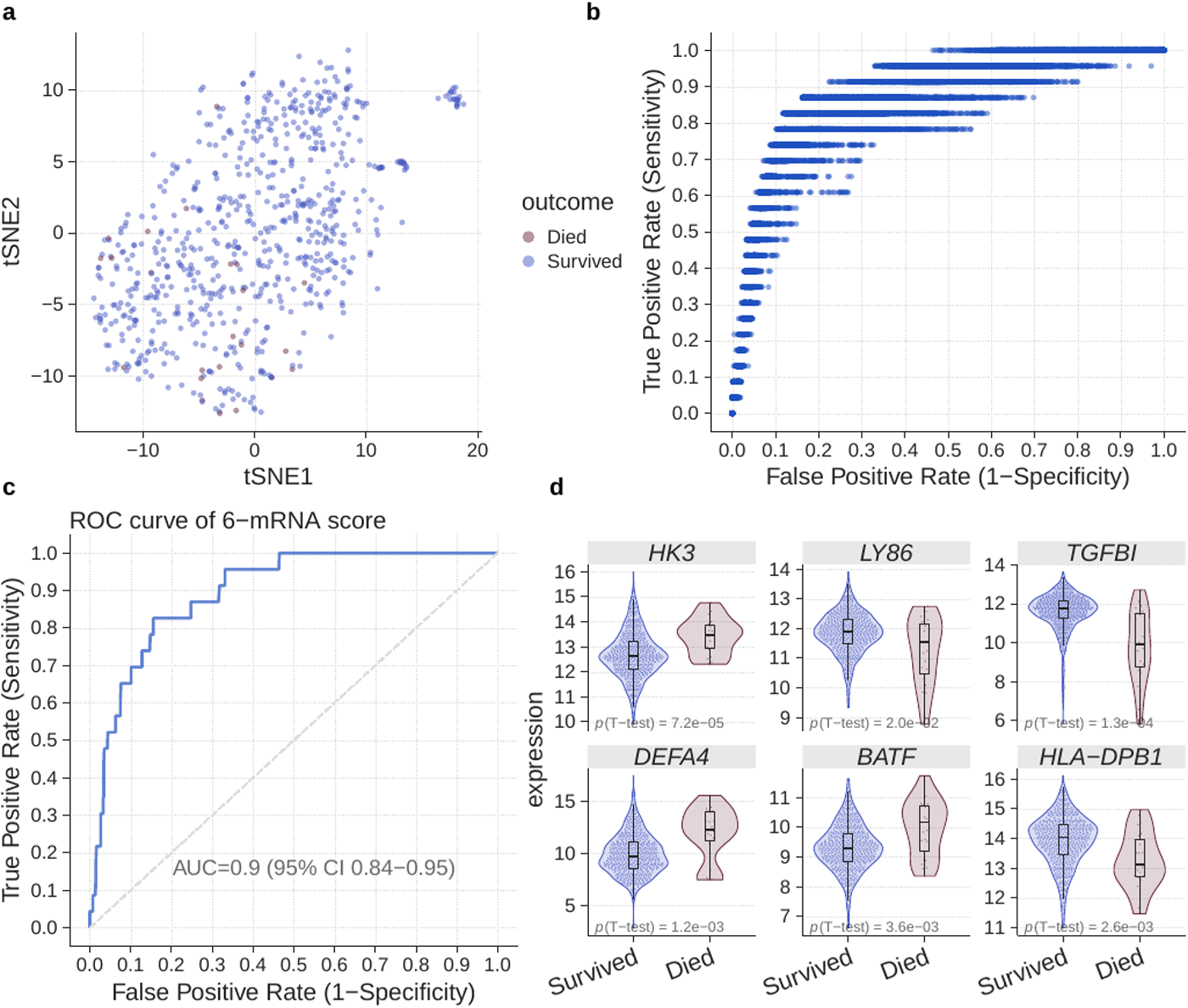
Training data for the 6-mRNA classifier. (a) Visualization of 705 samples across 21 studies in low dimension using t-SNE. (b) Logistic regression model selection. Each dot corresponds to a model defined by a combination of logistic regression hyperparameters and a decision threshold. Entire search space (100 hyperparameter configurations) is shown. (c) ROC plot for the best model. The plot is constructed using pooled probabilities from cross-validation folds. (d) Expression of the 6 genes used in the logistic regression model according to mortality outcomes.

### 6-mRNA logistic regression-based model accurately predicts viral patient mortality across multiple retrospective studies

Across the linear machine learning algorithms employed in our analyses, models using logistic regression had the highest mean AUROC for identifying patients with viral infection who died. Further, within logistic regression models, those trained using random cross-validation were more accurate than those trained using other variants of cross-validation. Finally, within the different 6-mRNA logistic regression-based models trained using CV, the model with highest AUROC used the following 6 genes: *TGFBI, DEFA4, LY86, BATF, HK3* and *HLA-DPB1*. It had an AUROC of 0.896 (95% CI: 0.844-0.949) (**Figures 2b and 2c; Supplementary Figure 1**). Each of the 6 genes were significantly differentially expressed between patients with viral infections who survived and those who did not, of which 3 genes (*DEFA4, BATF, HK3*) were higher and 3 genes (*TGFBI, LY86, HLA-DPB1*) were lower in those who died (**Figure 2d**). Based on the cross-validation, the 6-mRNA logistic regression model had a 91% sensitivity and 68% specificity for distinguishing patients with viral infection who died from those who survived. We used this model, referred to as the 6-mRNA classifier, as-is for validation in multiple independent retropective cohorts and a prospective cohort.

### 6-mRNA classifier is an age-independent predictor of mortality in patients with viral infections

Age is a known significant predictor of 30-day mortality in patients with respiratory viral infections. To assess the added value of the new prognostic information of the 6-mRNA classifier with regards to age in the training data, we fit a binary logistic regression model with age and pooled cross-validation 6-mRNA classifier probabilities as independent variables. The 6-mRNA score was significantly associated with increased risk of 30-day mortality (P<0.001), but age was not (P=0.06).

### Validation of the 6-mRNA classifier in multiple independent retrospective cohorts

We applied the locked 6-mRNA classifier to 1,417 transcriptome profiles of blood samples across 21 independent cohorts from patients with viral infections (663 healthy controls, 674 non-severe, 71 severe, 7 fatal) in 10 countries (**Supplementary Table 1**). Visualization of the 1,417 samples using expression of the 6 genes showed patients with severe outcome clustered closer (**Figure 3a**). Among the 6 genes, over-expressed genes (*HK3, DEFA4, BATF*) were positively correlated with severity of viral infection, and under-expressed gene (*HLA-DPB1, LY86, TGFBI*) were negatively correlated with severity (**Figure 3b**). Importantly, the 6-mRNA classifier score was positively correlated with severity and was significantly higher in patients with severe or fatal viral infection than those with non-severe viral infections or healthy controls (**Figure 3c**). Finally, the 6-mRNA classifier score distinguished patients with severe viral infection from those with non-severe viral infection (AUROC=0.91, 95% CI: 0.881-0.938) and healthy controls (AUROC=0.998, 95% CI: 0.994-1) (**Figure 3d**).

**Figure 3.**
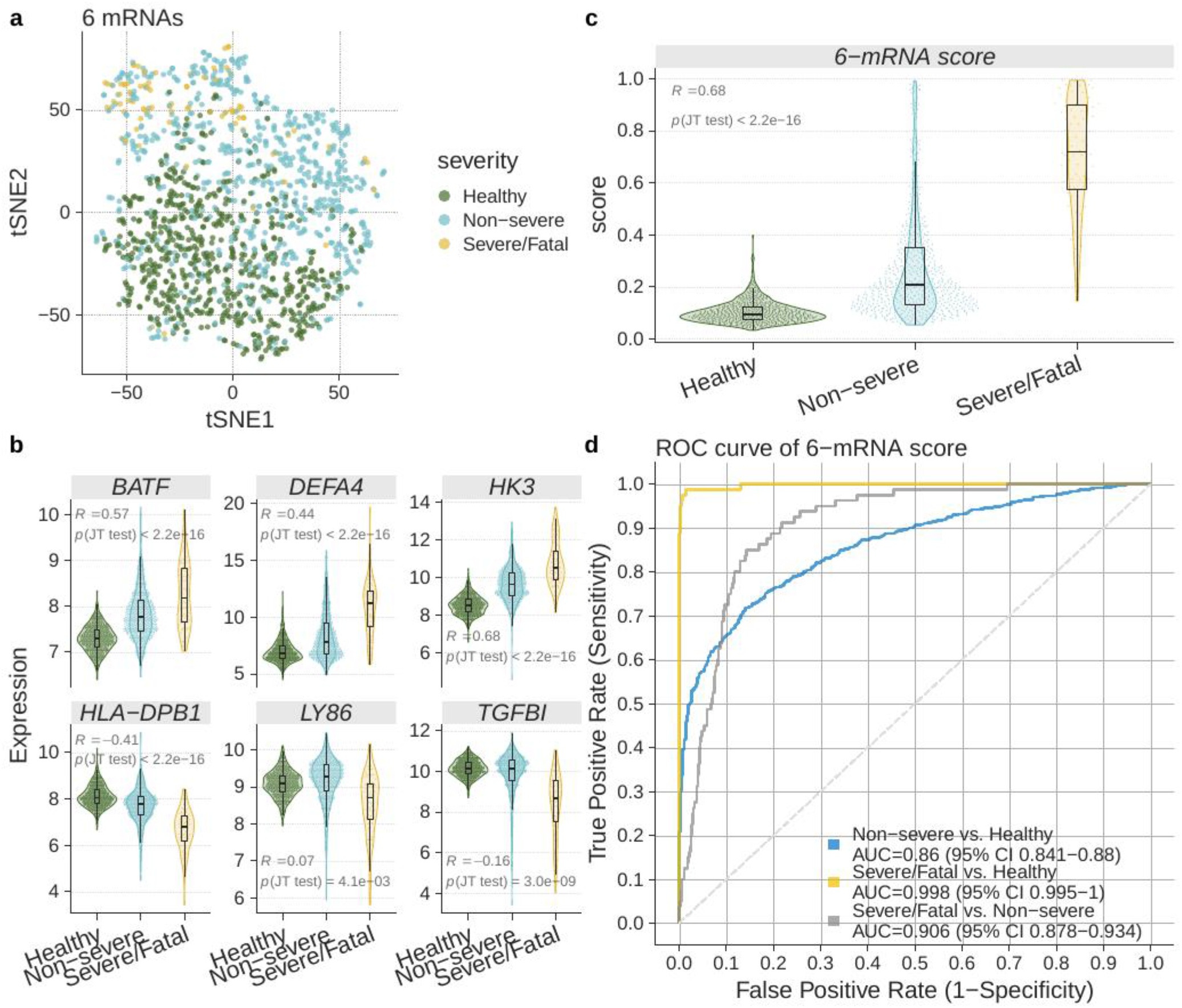
Validation of the 6-mRNA classifier in the independent retrospective non-COVID-19 cohorts. (a) Visualization of the samples using t-SNE. (b) Expression of the 6 genes used in the logistic regression model in patients with clinically relevant subgroups. (c) 6-mRNA classifier accurately distinguishes non-severe and severe patients with COVID-19 as well as those who died. (d) ROC plot for the subgroups.

We plotted ROC curves to assess the discriminative ability of the 6-mRNA classifier among the following subgroups of clinical interest: healthy controls, non-severe cases, severe, and fatal outcomes (**Fig. 3d**). Healthy controls are presented (though not mixed with non-severe viral infections in comparison) since some viral infections such as COVID-19 can be asymptomatic. All pairwise comparisons showed robust performance of the classifier on the independent data, achieving AUROC point-estimates between 0.86 (non-severe vs. healthy) and 1 (severe vs. healthy).

### Validation of the 6-mRNA logistic regression model in two independent COVID-19 cohorts

We further validated the 6-mRNA logistic regression model in two independent cohorts of patients with COVID-19. In one of the cohorts, we prospectively enrolled 97 adult patients with pneumonia by SARS-CoV-2 in Greece (Greece cohort). There were 47 patients with non-severe COVID-19 disease, whereas 50 had severe COVID-19, of which 16 died (**Table 2)**. We also used gene expression data for 100 COVID-19-positive participants in the GSE157103 study in Gene Expression Omnibus database (Albany cohort). There were 43 patients with non-severe COVID-19 disease and 57 with severe COVID-19. Visualization of both cohorts in low dimension using expression of the 6 mRNAs (without the classifier) revealed a degree of separation between patients with severe COVID-19 disease and those with non-severe disease (**Figure 4a**). When comparing expression of the 6 mRNAs in patients with non-severe COVID-19 disease to those with severe disease, expression of each changed statistically significant in the same direction as the training data in both cohorts (P<0.05) (**Figure 4b**).

**Figure 4.**
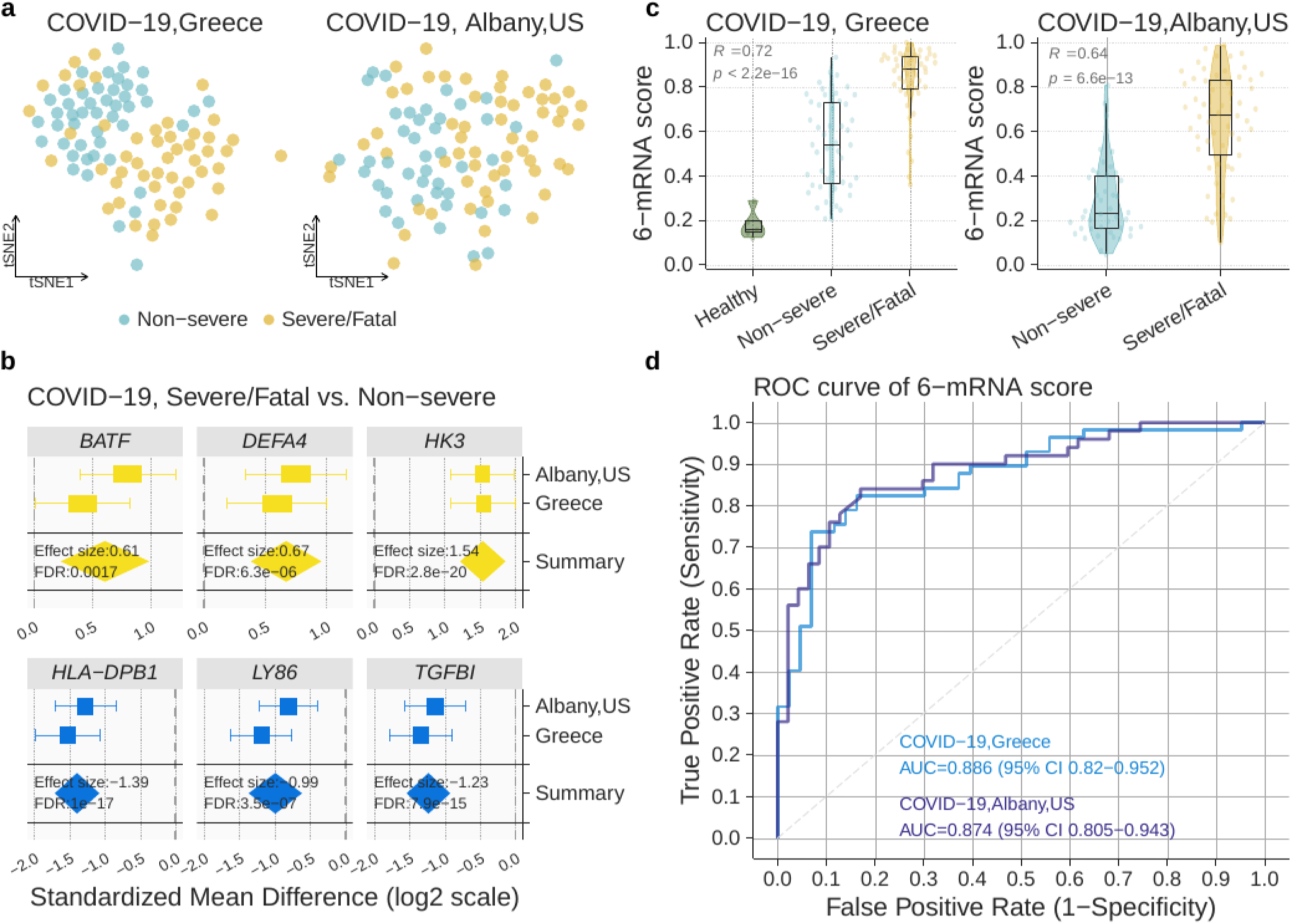
Validation of the 6-mRNA classifier in the COVID-19 cohorts. (A) Visualization of prospective (Greece, N=97) and retrospective (Albany, US, N=100) samples in the independent validation cohorts using t-SNE. (B) Expression of the 6 genes used in the logistic regression model in patients with severe/fatal and non-severe SARS-CoV-2 viral infection. (C) 6-mRNA classifier accurately distinguishes non-severe and severe patients with COVID-19 as well as those who died. (D) ROC plot for non-severe COVID-19 vs. severe or death (samples from healthy controls not included).

We applied the locked 6-mRNA classifier to the 97 COVID-19 patients and the 5 healthy controls in the Greece cohort. The 6-mRNA score was correlated with severity in both cohorts (Greece cohort: R=0.72, p<2.2e-16; Albany cohort: R=0.64, p=6.6e-13) (**Figure 4c**). In particular, the model distinguished patients with severe respiratory failure from non-severe patients with an AUROC of 0.89 (95% CI: 0.82-0.95) in the Greece cohort, and 0.87 (95% CI: 0.80-0.94) in the Albany cohort (**Figure 4d**).

We also assessed whether the 6-mRNA score is an independent predictor of severity in patients with COVID-19 by including other predictors of severity (age, SOFA score, CRP, PCT, lactate, and gender) in a logistic regression model. As expected, due to small sample size, and correlations between markers, no markers except SOFA were statistically significant predictors of severe respiratory failure (**Supplementary Table 3**).

For clinical applications, AUROC is a more relevant indicator of marker performance. To that end, we compared the 6-mRNA score to other clinical parameters of severity using AUROC (**Table 3 and Supplementary Figs. 2-3)**. The 6-mRNA score was the most accurate predictor of severe respiratory failure and death except SOFA. The AUROC confidence intervals were overlapping because the study was not powered to detect statistically significant differences. Of note, the 6-mRNA score was significantly more accurate for predicting severe respiratory failure and death than the only assay under the FDA Emergency Use Authorization, the IL-6. As a proxy for assessing how the 6-mRNA score might add to a clinician’s bedside severity assessment, we evaluated whether a combination of our classifier with the SOFA score improves over SOFA alone for the prediction of severe respiratory failure. The two scores together had an AUROC of 0.95; the continuous net reclassification improvement (cNRI) was 0.43 [95% CI: 0.04–0.81, P=0.03]. Together, these results suggest a potential improvement in clinical risk prediction when adding the 6-mRNA score to standard risk predictors; but definitive conclusion requires validation in additional independent data.

### Pooled results

We combined the predicted probabilities from the COVID-19 and non-COVID-19 independent cohorts and plotted the corresponding ROC graph (Fig. 5). The corresponding AUROC was 0.90 (95% CI: 0.87-0.92).

**Figure 5.**
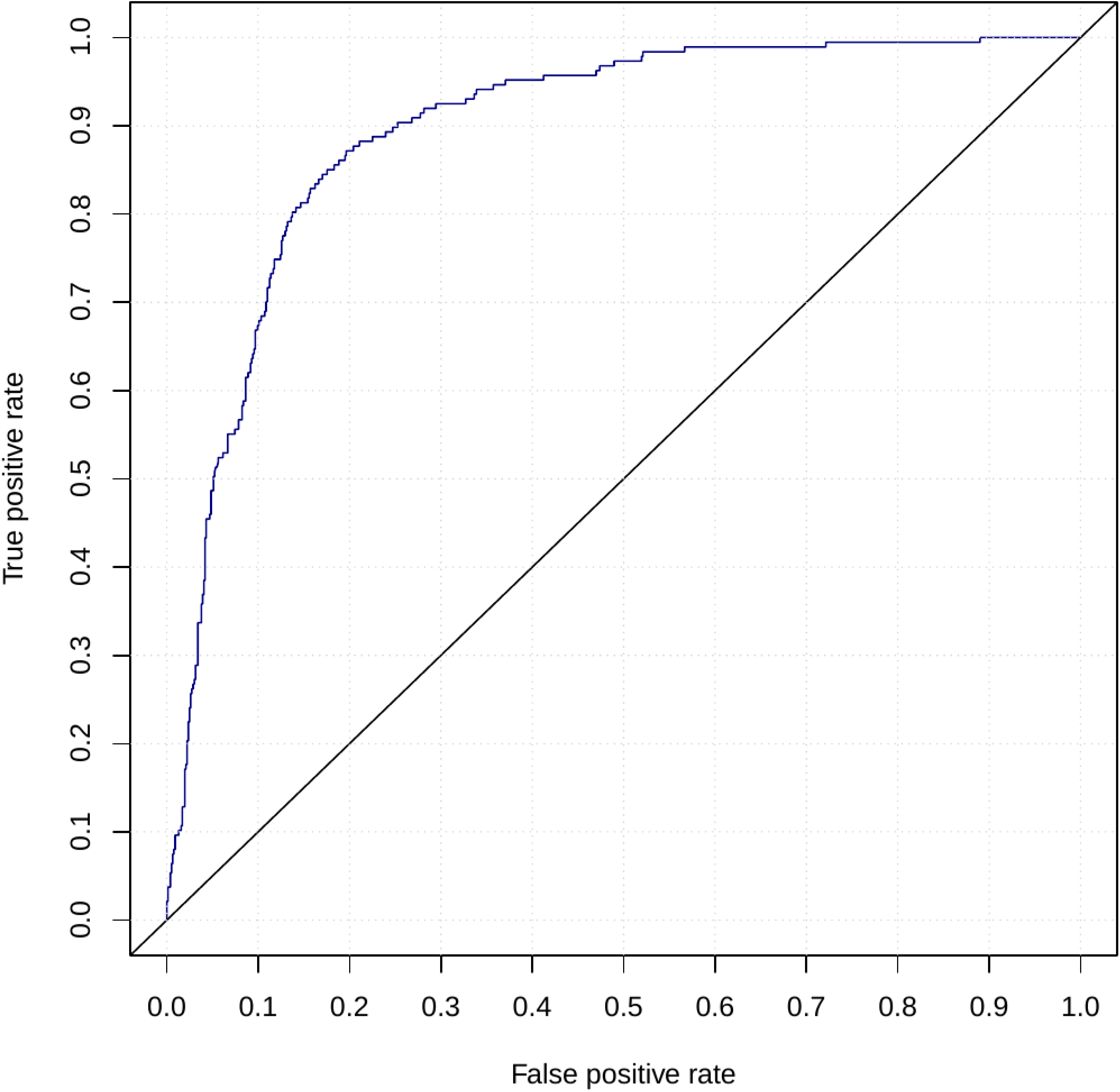
Validation of the 6-mRNA classifier in the COVID-19 and non-COVID-19 cohorts (pooled results of data in Figs. 3-4, excluding healthy subjects). The total number of samples was 951. The number of cases was 187. AUROC = 0.90 (95% CI: 0.87-0.92).

### Translation to a clinical report

To improve utility and adoption, a risk prediction score should be presented to clinicians in an intuitive and actionable test report. To that end, we discretized the 6-mRNA score in three bands: low-risk, intermediate-risk, and high-risk of severe outcome. The performance characteristics of each band are shown in **Table 4**. The table shows performance of the test on retrospective data (excluding healthy controls) using two versions of decision thresholds: thresholds optimized on the training data (**Table 4a**), and thresholds optimized using the retrospective test set (**Table 4b**). The outcome was severe infection. **Tables 4c, d** show corresponding results on the COVID-19 data, using severe respiratory failure as outcome.

**Table 4.**
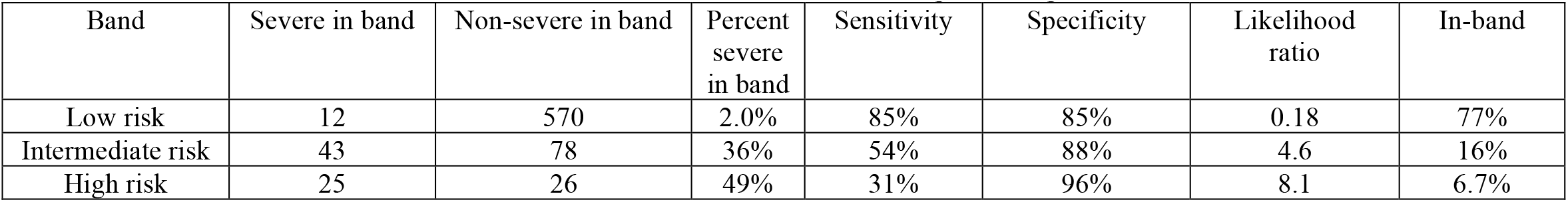
Test characteristics of the 6-mRNA score in non-COVID-19 and COVID-19 patients using the three-band test report. “Severe in band” is the number of patients with severe viral infection assigned to the corresponding band. “Non-severe in band” is the number of patients with non-severe viral infection assigned to the corresponding band. The “Percent severe in band” is the percentage of patients in the band who had severe outcome. The “In-band” column is the percentage of patients assigned by the classifier to the corresponding band. **Table 4a**. non-COVID-19 results. The band thresholds were set using training data and locked.

**Table 4b.** non-COVID-19 results. The band thresholds were set using the retrospective data.

| Band | Severe in band | Non-severe in band | Percent severe in band | Sensitivity | Specificity | Likelihood ratio | In-band |
| --- | --- | --- | --- | --- | --- | --- | --- |
| Low risk | 2 | 420 | 0.47% | 98% | 62% | 0.04 | 56% |
| Intermediate risk | 68 | 246 | 22% | 85% | 64% | 2.3 | 42% |
| High risk | 10 | 8 | 56% | 12% | 99% | 11 | 2.4% |

**Table 4c.** Prospective COVID-19 results. The band thresholds were set using training data and locked.

| Band | Severe in band | Non-severe in band | Percent severe in band | Sensitivity | Specificity | Likelihood ratio | In-band |
| --- | --- | --- | --- | --- | --- | --- | --- |
| Low risk | 2 | 18 | 10% | 96% | 38% | 0.10 | 21% |
| Intermediate risk | 20 | 28 | 42% | 40% | 40% | 0.67 | 50% |
| High risk | 28 | 1 | 97% | 56% | 98% | 26 | 30% |

**Table 4d.**
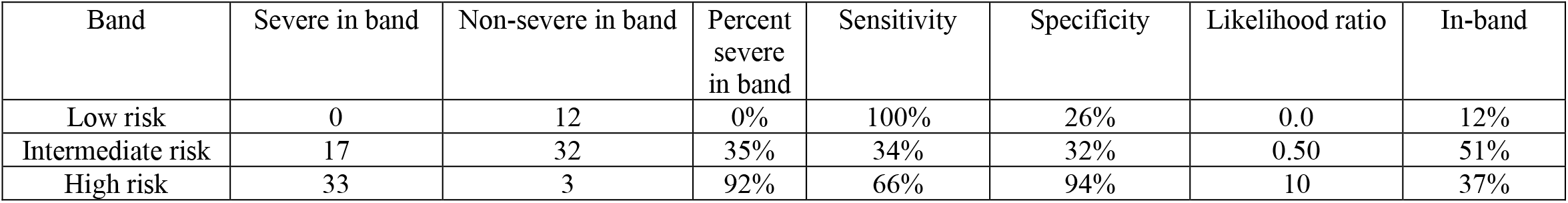
Prospective COVID-19 results. The band thresholds were set using the prospective data.

| Band | Severe in band | Non-severe in band | Percent severe in band | Sensitivity | Specificity | Likelihood ratio | In-band |
| --- | --- | --- | --- | --- | --- | --- | --- |
| Low risk | 0 | 12 | 0% | 100% | 26% | 0.0 | 12% |
| Intermediate risk | 17 | 32 | 35% | 34% | 32% | 0.50 | 51% |
| High risk | 33 | 3 | 92% | 66% | 94% | 10 | 37% |

**Table 4e.** Retrospective COVID-19 results. The band thresholds were set using training data and locked.

| Band | Severe in band | Non-severe in band | Percent severe in band | Sensitivity | Specificity | Likelihood ratio | In-band |
| --- | --- | --- | --- | --- | --- | --- | --- |
| Low risk | 14 | 37 | 27% | 75% | 86% | 0.28 | 51% |
| Intermediate risk | 33 | 6 | 85% | 58% | 86% | 4.1 | 39% |
| High risk | 10 | 0 | 100% | 18% | 100% | Inf | 10% |

**Table 4f.** Retrospective COVID-19 results. The band thresholds were set using the retrospective data.

| Band | Severe in band | Non-severe in band | Percent severe in band | Sensitivity | Specificity | Likelihood ratio | In-band |
| --- | --- | --- | --- | --- | --- | --- | --- |
| Low risk | 1 | 16 | 6% | 98% | 37% | 0.047 | 17% |
| Intermediate risk | 14 | 24 | 37% | 25% | 44% | 0.44 | 38% |
| High risk | 42 | 3 | 93% | 74% | 93% | 11 | 45% |

### Translation to a rapid assay

Any risk prediction score should be rapid enough to fit into clinical workflows. We thus developed a LAMP assay as a proof of concept for a rapid 6-mRNA test. We further showed that across 61 clinical samples from healthy controls and acute infections of varying severities that the LAMP 6-mRNA score and the reference NanoString 6-mRNA score had very high correlation (r=0.95; **Supplementary Figure 4)**. These results demonstrate that with further optimization the 6-mRNA model could be translated into a clinical assay to run in less than 30 minutes.

## Discussion

The severe economic and societal cost of the ongoing COVID-19 pandemic, the fourth viral pandemic since 2009, has underscored the urgent need for a prognostic test that can help stratify patients as to who can safely convalesce at home in isolation and who needs to be monitored closely. Here we integrated 705 peripheral blood transcriptome profiles across 21 heterogeneous studies from patients with viral infections, none of whom were infected with SARS-CoV-2. Despite the substantial biological, clinical, and technical heterogeneity across these studies, we identified a 6-mRNA host-response signature that distinguished patients with severe viral infections from those without. We demonstrated generalizability of this 6-mRNA model first in a set of 21 independent heterogeneous cohorts of 1,417 retrospectively profiled samples, and then in two independent retrospectively and prospectively collected cohorts of patients with SARS-CoV-2 infection, in United States and Greece, respectively. In each validation analysis, the 6-mRNA classifier accurately distinguished patients with severe outcome from those with non-severe outcomes, irrespective of the infecting virus, including SAR-CoV-2. Importantly, across each analysis, the 6-mRNA classifier had similar accuracy, measured by AUROC, demonstrating its generalizability and robustness to biological, clinical, and technical heterogeneity. Although this study was focused on development of a clinical tool, not a description of transcriptome-wide changes, the applicability of the signature across viral infections further demonstrates that host factors associated with severe outcomes are conserved across viral infections, which is in line with our recent large-scale analysis (*20*).

While many risk-stratification scores and biomarkers exist, few are focused specifically on viral infections. Of the recent models specifically designed for COVID-19, most are trained and validated in the same homogenous cohorts, and their generalizability to other viruses is unknown because they have not been tested across other viral infections (*14*). Consequently, when a new virus, such as SARS-CoV-2, emerges, their utility is substantially limited. However, we have repeatedly demonstrated that the host response to viral infections is conserved and distinct from the host response to other acute conditions *(15-20)*.

Here, building upon our prior results, we developed a 6-mRNA classifier specifically trained in patients with viral infection to risk stratify better than other existing biomarkers. Further, the only assay authorized for clinical use in risk-stratifying COVID-19 (IL-6 measured in blood), substantially underperformed our proposed 6-mRNA model here. That said, the nominal improvement over existing biomarkers (**Table 3**) for prediction of severe respiratory failure requires larger cohorts to confirm statistical significance. The 6-mRNA score is nominally worse than SOFA, but SOFA requires 24 hours to calculate, while the 6-mRNA score could be run in 30 minutes, demonstrating its utility as a triage test. The synergy (positive NRI) in combination with SOFA also suggests that the 6-mRNA score could improve practice in combination with clinical gestalt. The 6-mRNA score has been reduced to practice as a rapid isothermal quantitative RT-LAMP assay, suggesting that it may be practical to implement in the clinic with further development.

Our goal in this study was not to investigate underlying biological mechanisms, but to address the urgent need for a prognostic test in SARS-CoV-2 pandemic, and to improve our preparedness for future pandemics. However, using immunoStates database (https://metasignature.khatrilab.stanford.edu) (*44*), we found 5 out of the 6 genes (*HK3, DEFA4, TGFBI, LY86, HLA-DPB1*) are highly expressed in myeloid cells, including monocytes, myeloid dendritic cells, and granulocytes. This is in line with our recent results demonstrating that myeloid cells are the primary source of conserved host response to viral infection (*20*). Further, we have previously found that *DEFA4* is over-expressed in patients with dengue virus infection who progress to severe infection (*44*), and in those with higher risk of mortality in patients with sepsis (*18*). *HLA-DPB1* belongs to the HLA class II beta chain paralogues, and plays a central role in the immune system by presenting peptides derived from extracellular proteins. Class II molecules are expressed in antigen presenting cells (B lymphocytes, dendritic cells, macrophages). Reduced expression of *HLA-DPB1* described herein is fully compatible with the decreased expression of HLA-DR on the cell membranes of circulating monocytes of patients with severe respiratory failure by SARS-CoV-2. This is a unique immune dysregulation where despite the down-regulation of HLA-DR monocytes remain potent for the production of pro-inflammatory cytokines, namely TNFα and IL-6. This complex immune dysregulation fully differentiates critically ill patients with COVID-19 from patients with bacterial sepsis (*46*) in patients with severe outcome and suggests dysfunctional antigen presentation that should be further investigated. Similarly, *BATF* is significantly over-expressed, and *TGFBI* is significantly under-expressed in patients with sepsis compared to those with systemic inflammatory response syndrome (SIRS) (*15*). Finally, lower expression of *TGFBI* and *LY86* in peripheral blood is associated with increased risk of mortality in patients with sepsis (*18*). These results further suggest that there may be a common underlying host immune response associated with severe outcome in infections, irrespective of bacterial or viral infection. Consistent differential expression of these genes in patients with a severe infectious disease across heterogeneous datasets lend further support to our hypothesis that dysregulation in host response can be leveraged to stratify patients in high- and low-risk groups.

Our study has several limitations. First, our study uses retrospective data with large amount of heterogeneity for discovery of the 6-mRNA signature; such heterogeneity could hide unknown confounders in classifier development. However, our successful representation of biological, clinical, and technical heterogeneity also increased the *a priori* odds of identifying a parsimonious set of generalizable prognostic biomarkers suitable for clinical translation as a point-of-care. Second, owing to practical considerations for urgent need, we focused on a preselected panel of mRNAs. It is possible that similar analysis using the whole transcriptome data would find additional signatures, though with less clinical data. Third, a common limitation in all these types of pandemic observational studies is a lack of understanding of the effect of time from symptoms onset. Finally, additional larger prospective cohorts are needed to further confirm the accuracy of the 6-mRNA model in distinguishing patients at high risk of progressing to severe outcomes from those who do not.

Overall, our results show that once translated into a rapid assay and validated in larger prospective cohorts, this 6-mRNA prognostic score could be used as a clinical tool to help triage patients after diagnosis with SARS-CoV-2 or other viral infections such as influenza. Improved triage could reduce morbidity and mortality while allocating resources more effectively. By identifying patients at high risk to develop severe viral infection, i.e., the group of patients with viral infection who will benefit the most from close observation and antiviral therapy, our 6-mRNA signature can also guide patient selection and possibly endpoint measurements in clinical trials aimed at evaluating emerging anti-viral therapies. This is particularly important in the setting of current COVID-19 pandemic, but also useful in future pandemics or even seasonal influenza.

## Conclusions

With further study, the classifier could assist in the risk assessment of COVID-19 and other acute viral infections patients to determine severity and level of care, thereby improving patient management and reducing healthcare burden.

## Data Availability

The public cohorts are available under their respective study IDs. The Stanford ICU Databank study is available at https://doi.org/10.1038/s41467-020-14975-w. The COVID-19 NanoString data are available upon reasonable request from the authors.

https://doi.org/10.1038/s41467-020-14975-w

## List of abbreviations

COVID-19; SARS-CoV-2; ARDS; CD3; CD4; ML; mRNA; RNAseq; IMX-BVN-2; CV; LOSO; AUROC; PBMC; ED; ICU; SOFA; HIV; cDNA; gDNA; LAMP; PCR; CI; FDA; cNRI

## Declarations

### Ethics approval and consent to participate

COVID-19 blood samples were collected between March and April 2020. The studies were conducted following approvals for the collection of biomaterial for transcriptomic analysis for patients with lower respiratory tract infections provided by the Ethics Committees of the participating hospitals. The studies were conducted under the 23/12.08.2019 approval of the Ethics Committee of Sotiria Athens General Hospital; and the 26.02.2019 approval of the Ethics Committee of ATTIKON University General Hospital. Written informed consent was provided by patients or by first-degree relatives in case of patients unable to consent. For Australia/WIMR study, the written consent is provided by all patients or by first-degree relatives in case of patients unable to consent. The study was approved by the human research ethics committee of the Nepean Blue Mountains Area Health Services. For PREVISE study, the IRB was approved by COMITE DE ETICA DE LA INVESTIGACION CON MEDICAMENTOS DEL AREA DE SALUD DE SALAMANCA, Paseo de San Vicente, 58-18, 237007 Salamanca, Spain. For healthy commercial controls, blood RNA tubes were prospectively collected from healthy controls (HC) through a commercial vendor (BioIVT) under IRB approval (Western IRB #2016165) using informed consent. Stanford ICU databank and PROMPT studies have previously been published and provided IRB details.

All necessary patient/participant consent has been obtained and the appropriate institutional forms have been archived.

### Consent for publication

All necessary patient/participant consent has been obtained and the appropriate institutional forms have been archived.

## Availability of data and materials

The public cohorts are available under their respective study IDs. The Stanford ICU Databank study is available at https://doi.org/10.1038/s41467-020-14975-w. The COVID-19 NanoString data are available upon reasonable request.

## Authors’ contributions

TES, LB, PK, and EJGB designed the study; BT, KL, WSK, MG, RS, MS, RA, JAN, SM, CH, NA, PK, MK, GD, OL conducted clinical studies; LB, HZ, JW, UM, HZ and RL performed data modelling and statistical analysis; YL, AMR, DD, JT, LMJ, MD collected and curated transcriptome data from public repositories; DR, MR, and SC built rapid assays and processed samples; LB, PK, OL, and TES wrote the manuscript; all authors critically revised and approved the manuscript.

## Acknowledgements

We are grateful for the expert support in sample shipment and receipt of Ashley Prasse Miller and Mario Esquivel. We thank Jesús Bermejo-Martin for assistance with the PREVISE clinical collaboration.

