## Supplementary information for "A 6-mRNA host response whole-blood classifier trained on pre-pandemic data accurately predicts severity in COVID-19 and other acute viral infections"

**Table of Contents:**

Supplementary Figures 1-4

Supplementary Tables 1-3

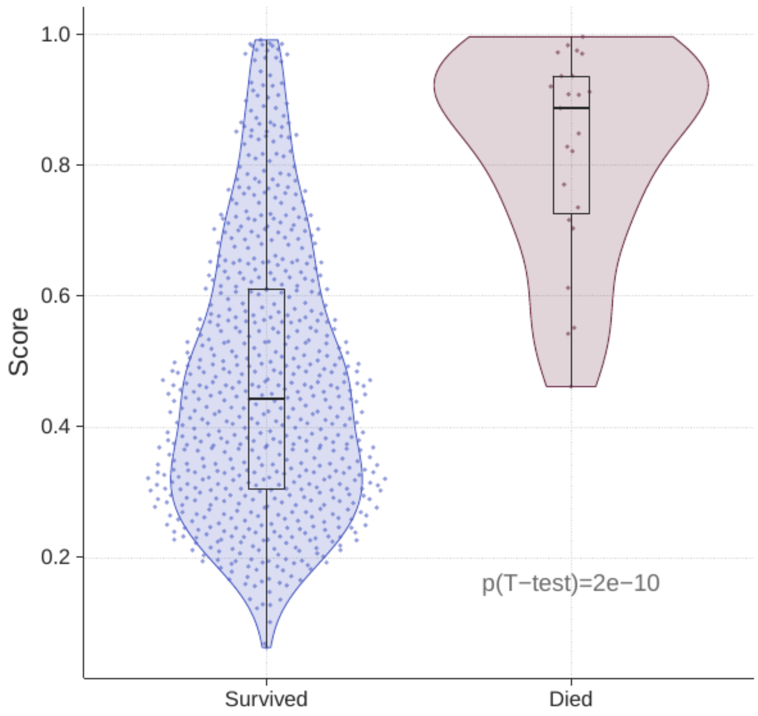


**Supplementary Figure 1.** Distribution of the pooled training set cross-validation 6-mRNA score for the best logistic regression model. Blue = survivors, red=non-survivors.

**Supplementary Figure 2.** Comparison of ROC curves for 6-mRNA ("CoVerity") classifier and clinical markers for severe respiratory failure outcome.

SOFA APACHE

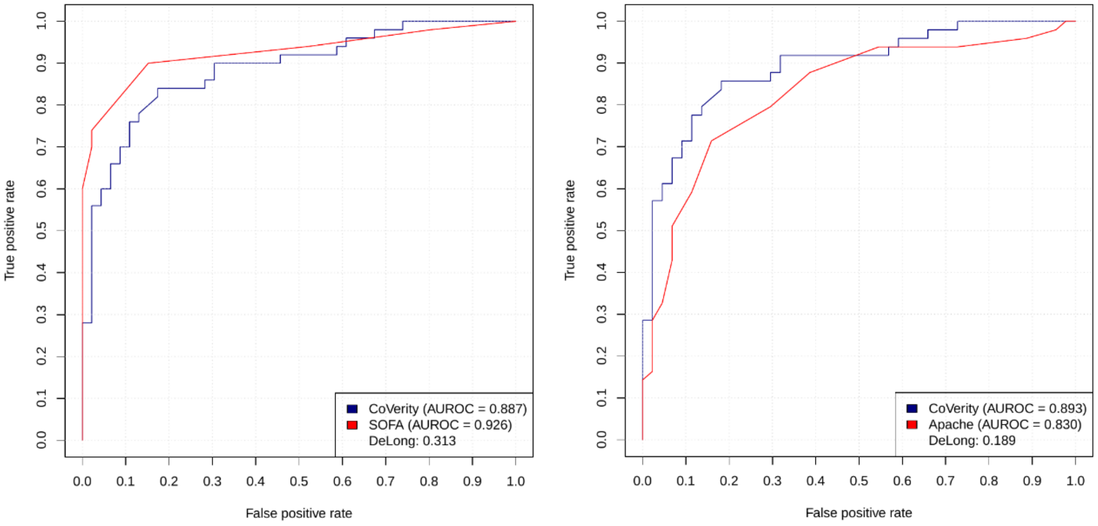

Age

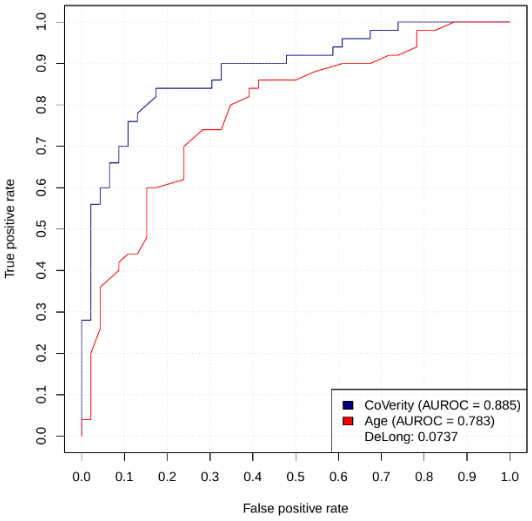


PCT

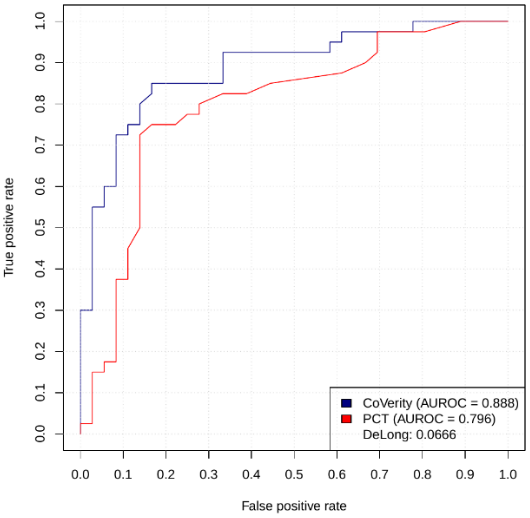


CRP

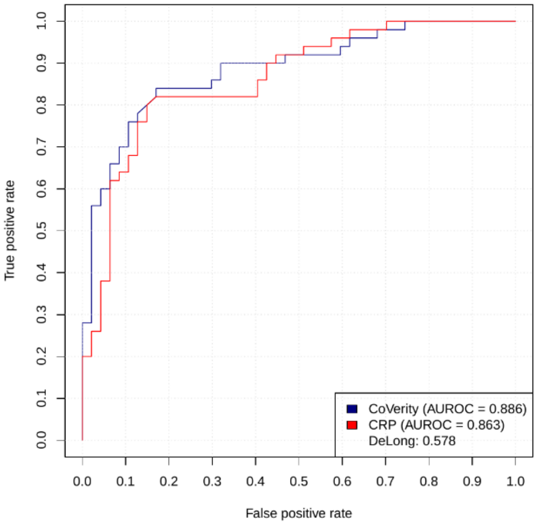


Lactate

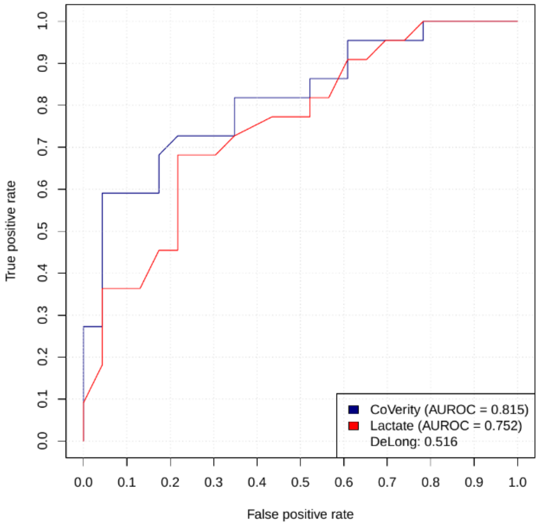

IL-6

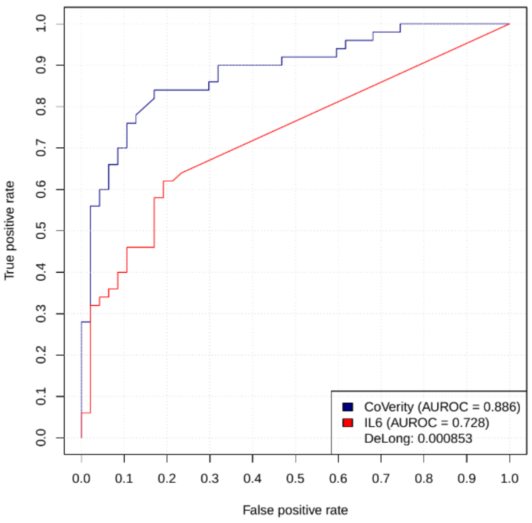


suPAR

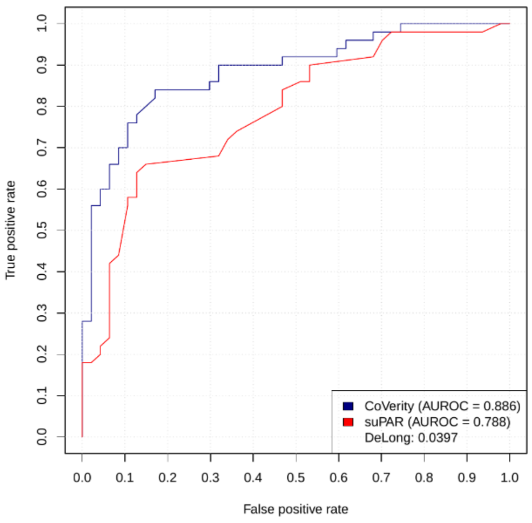


**Supplementary Figure 3.** Comparison of ROC curves for 6-mRNA classifier ("CoVerity") and clinical markers for death outcome.

SOFA

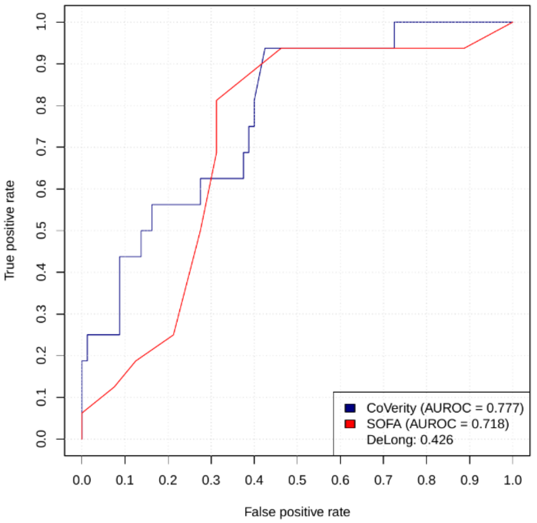


APACHE

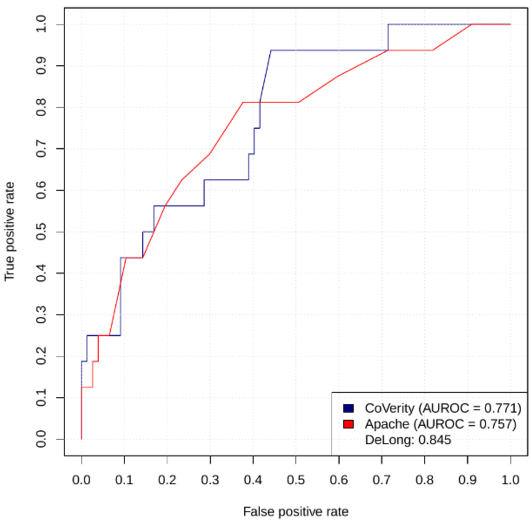

Age

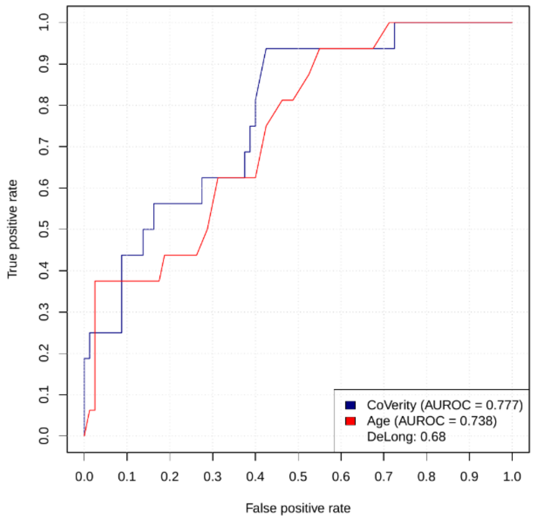


PCT

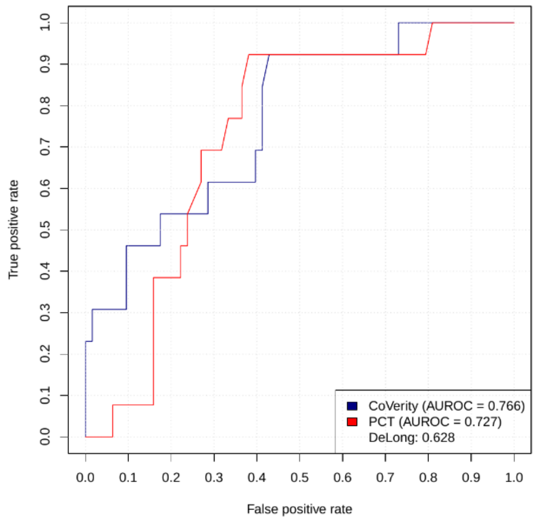


CRP

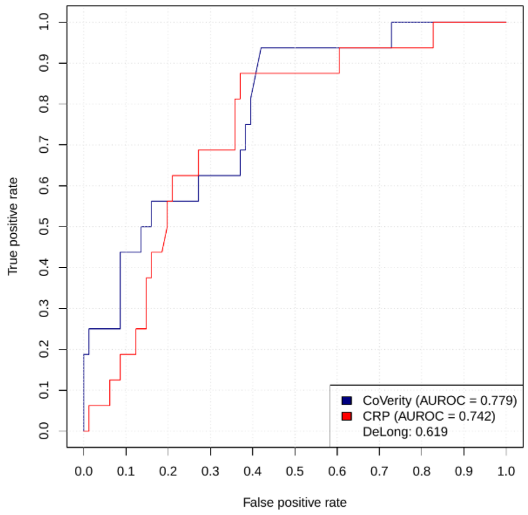


Lactate

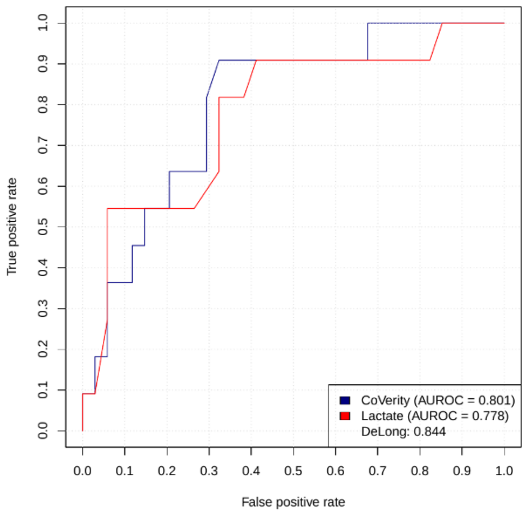


IL-6

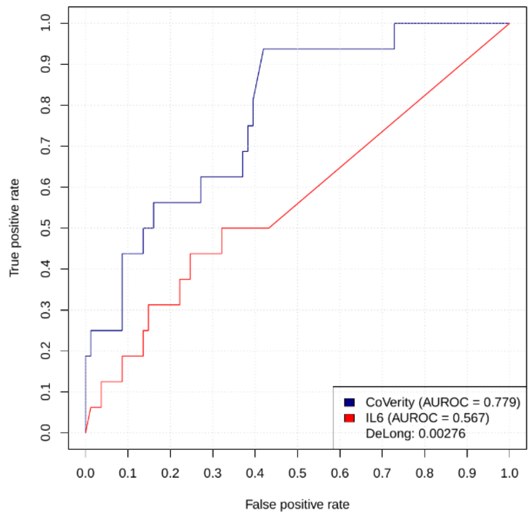


suPAR

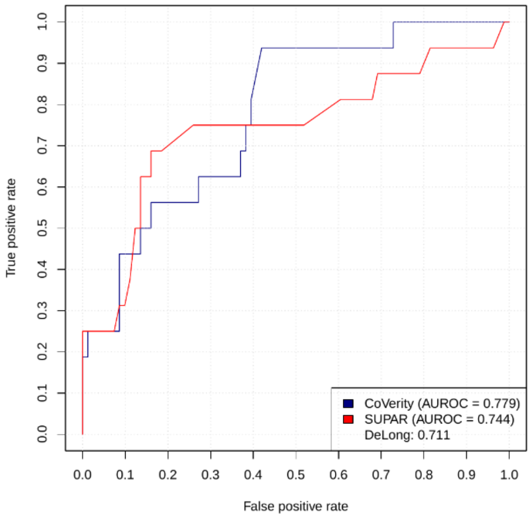

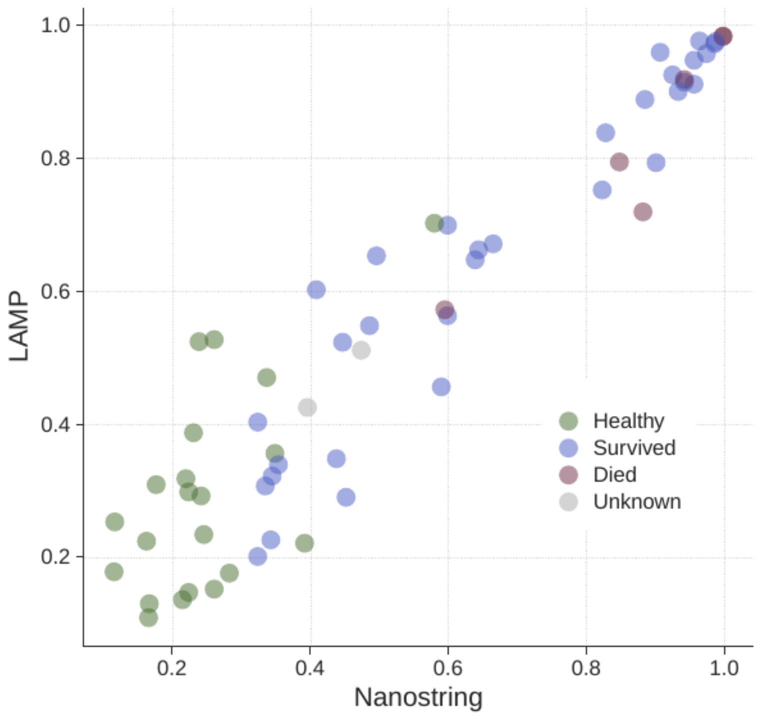


**Supplementary Figure 4.** Correlation of the 6-mRNA classifier scores using rapid qRT-LAMP panel and NanoString nCounter gold standard shows excellent agreement (Pearson R = 0.95) across n=61 clinical samples.

**Supplementary Table 1.** Characteristics of retrospective viral infection (non-COVID-19) studies used for independent validation.

| Study identifier | First author or PI | Study description | Timing of sample collection | N (total/healthy/non-severe/severe/fatal) | Age | Male (n (%)) | Country | Platform |
|---|---|---|---|---|---|---|---|---|
| GSE103842 | Rodriguez-Fernandez | RSV infected infants | Within 24 hours of hospitalization | 74,12,62,0,0 | Child (0-2 years) | 48(65) | USA | Illumina |
| GSE111368 | Dunning | Patients with severe influenza with or without bacterial co-infection | Samples were obtained at three time points: T1 (recruitment), T2 (approximately 48 h after T1) and T3 (at least 4 weeks after T1) | 239,130,81,28,0 | Adult (18-71 years) | 111(46) | UK | Illumina |
| GSE20346 | Parnell | Adults with CAP | Hospital admission | 22,18,0,4,0 | Adult (21-75 years) | 7(32) | Australia | Illumina |
| GSE27131 | Berdal | Patients with documented influenza, bilateral chest infiltrates, and in need of ventilation support, without significant co-morbidity | Admission to ICU | 13,7,0,3,3 | Adult (25-59 years) | 9(69) | Norway | Affymetrix |

| | | | | | | | | |
|---|---|---|---|---|---|---|---|---|
| GSE77087 | de Steenhuijsen | Outpatient and inpatient RSV patients | Either at the outpatient clinics or within a median of 24 hours of admission in the pediatric ward or the pediatric ICU | 104,23,81,0,0 | Child (0-2 years) | 67(64) | USA | Illumina |
| GSE67059 | Heinonen | Asymptomatic and symptomatic rhinovirus in children | ED (outpatients) or within 48 hours of hospitalization (inpatients) | 137,37,100,0,0 | Child (0-2 years) | 87(64) | USA, Finland, Spain | Illumina |
| GSE21802 | Bermejo-Martin | Patients attending to the participants ICUs with primary viral pneumonia during the acute phase of influenza virus illness with acute respiratory distress and unequivocal alveolar opacification involving two or more lobes with negative respiratory and blood bacterial cultures at admission | Admission to ICU | 20,4,12,2,2 | Adult (18-65 years) | 12(60) | Spain | Illumina |
| GSE66099 | Sweeney, Alder | Septic children in PICU | Admission to ICU | 58,47,0,9,2 | Child (0-10 years) | 32(55) | USA | Affymetrix |
| GSE101702 | Tang, Zerbib | Influenza patients with varying severity of infection | Within 24 hours of their presentation to the hospital | 159,52,107,0,0 | Adult (17-90 years) | 63(40) | Australia, Canada, Germany | Agilent |
| GSE17156_FLU | Zaas | Influenza challenge study | Multiple time points | 25,17,8,0,0 | Adult (>18 years) | 12(48) | USA, UK | Affymetrix |
| GSE17156_RSV | Zaas | RSV challenge study | Multiple time points | 29,20,9,0,0 | Adult (>18 years) | 16(55) | USA, UK | Affymetrix |
| GSE17156_RHINO | Zaas | Rhinovirus challenge study | Multiple time points | 29,19,10,0,0 | Adult (>18 years) | 16(55) | USA, UK | Affymetrix |
| GSE40012 | Parnell | Adults with CAP | Within 24 hours of admission to ICU | 38,36,0,2,0 | Adult (22-75 years) | 13(34) | Australia, Hong Kong | Illumina |
| GSE68004 | Jaggi | Kawasaki disease compared to other febrile patients | Hospital admission | 56,37,19,0,0 | Child (0-16 years) | 25(45) | USA | Illumina |
| EMTAB5195 | Jong | Respiratory syncytial virus infected infants | Within 24 hours of presentation to the hospital | 43,4,21,18,0 | Child (0-2 years) | 27(63) | Netherlands | Affymetrix |
| GSE6269 | Ramilo | Sepsis patients with influenza or bacterial infection | | 24,6,18,0,0 | Child (0-18 years) | 15(62) | USA | Affymetrix, Illumina |
| GSE68310 | Zhai | Influenza and other acute respiratory viral infections | Multiple time points | 157,128,29,0,0 | Adult (18-49) | 77(49) | USA | Illumina |
| GSE117827 | Yu | Children with acute viral infection | Hospital admission | 19,6,13,0,0 | Child (0-11 years) | 14(74) | USA | Affymetrix |
| GSE25504 | Smith, Dickinson | Septic neonates | Hospital admission, at the time of first clinical signs of suspected sepsis | 9,6,3,0,0 | Child (0-1 year) | 9(100) | UK | Affymetrix |

| | | | | | | | | |
|---|---|---|---|---|---|---|---|---|
| GSE4607 | Wong, Cvijanovich | Septic children in PICU | Within 24 hours of admission to ICU | 22,15,0,5,2 | Child (0-10 years) | 14(64) | USA | Affymetrix |
| GSE38900 | Mejias | Children with acute LRTI | Hospital admission | 140,39,101,0,0 | Child (0-2 years) | 76(54) | USA, Finland | Illumina |

**Supplementary Table 2:** Oligonucleotide sequences for detection of 6 informative viral severity markers.

| **Oligo ID** | **Sequence** |
|---|---|
| PD HK3v4 F3 | ACCTGAGGAGAGTGACTAGCTTCT |
| PD HK3v4 B3 | GCCTGCTCCATGGAACCCAAGA |
| PD HK3v4 FIP | TCAGAGCAACTCAGGGTTTCTTCCCCACTGTGGAAGCTCATGGAC |
| PD HK3v4 BIP | TCAGAGCTGGTGCAGGAGTGCGCTGGCTTGGATCTGCTGTAGC |
| PD HK3v4 FL | CCGCAACCCTGAAGACCCA |
| PD HK3v4 BL | GCAGTTCAAGGTGACAAGGGCAC |
| PD BATFv3 F3 | CTGAGTGTGAGAGCCCGGAAGATTT |
| PD BATFv3 B3 | TGTTCAGCACCGACGTGAAGTACTT |
| PD BATFv3 FIP | TACGATTTTTCTCCCTCCTCTGAACTCTTCAGCAGTGACTCCAGCTTCAGC |
| PD BATFv3 BIP | GAAGAGCCGACAGAGGCAGTGCTTGATCTCCTTGCGTAGAGCC |
| PD BATFv3 LF | CATCAGATGAGTCCTGTTTGCCAGG |
| PD BATFv3 LB | GCACCTGGAGAGCGAAGACCT |
| PE DEFA4 i2v4-12 F3 | AGGTGATGAGGCTCCAGG |
| PE DEFA4 i2v4-12 B3 | TGAAACTCACACCACCAATGA |
| PE DEFA4 i2v4-12 FIP | ACCTGAAGAGCAGAGCTTTTATCCCAGCGTGGGCCAGAAGAC |
| PE DEFA4 i2v4-12 BIP | TCAGGCTCAACAAGGGGCATGGCAGTTCCCAACACGAAGTT |
| PE DEFA4 i2v4-12 FL | GCTCTTGCAGATTAGTATTCTGCCGG |
| PE DEFA4 i2v4-12 BL | GTCCTGTATAGATAAAGGAAACGTA |
| PD LY86v9 F3 | CTTGACCTAGCTCTCATGTCTCAA |
| PD LY86v9 B3 | CACATGATAGTAGCATTGGCACA |
| PD LY86v9 FIP | GCATAGTAAATCTGCTCTCCTTTCCGGCTCATCTGTTTTGAATTTCTCCTA |
| PD LY86v9 BIP | GGCCTGTCAATAATCCTGAATTTACTGGTGGACCGTTTTTCAGTGTAC |
| PD LY86v9 FL | CCACAGAAAGAAAACTTGGGCA |
| PD LY86v9 BL | CCTCAGGGAGAATACCAGGTTT |
| PD TGFBIv4 F3 | GGTGATGAAATCCTGGTTAGCGGA |
| PD TGFBIv4 B3 | CGCTGATGCTTGTTTGAAGATCTC |
| PD TGFBIv4 FIP | AGGCTCCTTGTTGACACTCACCACGCCCTGGTGCGGCTAAAGTCT |
| PD TGFBIv4 BIP | TGACATCATGGCCACAAATGGCGTCAGAGTCTGCAAGTTCATCCCCT |
| PD TGFBIv4 LF | GCTGACTTCCAGCTTGTCACCT |
| PD TGFBIv4 LB | CTCCAGCCAACAGACCTCAGGAA |
| PE HLA-DPB1v1 F3 | CTGCGGAGTACTGGAACAG |
| PE HLA-DPB1v1 B3 | CGTCACGTGGCAGACAAG |
| PE HLA-DPB1v1 FIP | GCCCAGCTCGTAGTTGTGTCTGGAAGGACATCCTGGAGGAGA |
| PE HLA-DPB1v1 BIP | CCGAGTCCAGCCTAGGGTGAGGTTGTGGTGCTGCAAGG |
| PE HLA-DPB1v1-1 FL | ATCCTGTCCGGCACTGC |

| | |
|---|---|
| PE HLA-DPB1v1-1 BL | ATGTTTCCCCCTCCAAGAAGG |

**Supplementary Table 3.** Multiple regression model in the COVID-19 cohort with severe respiratory failure as the dependent variable.

| | Estimate | Std. Error | Statistic | P-value |
|---|---|---|---|---|
| (Intercept) | -13.5 | 4.36 | -3.10 | **0.00197** |
| 6-mRNA score | 5.42 | 4.04 | 1.34 | 0.181 |
| Age (years) | 0.104 | 0.0460 | 2.26 | 0.0239 |
| CRP (mg/l) | 0.0132 | 0.00782 | 1.70 | 0.090 |
| PCT (ng/ml) | -0.185 | 0.210 | -0.882 | 0.378 |
| Gender (Male) | -1.37 | 1.297 | -1.06 | 0.290 |
| SOFA | 0.73 | 0.301 | 2.42 | **0.016** |
